# Detection of severe acute respiratory syndrome coronavirus 2 (SARS-CoV-2) in a fourplex real-time quantitative reverse transcription-PCR assays

**DOI:** 10.1101/2021.03.23.21254196

**Authors:** Mathieu Durand, Philippe Thibault, Simon Lévesque, Ariane Brault, Alex Carignan, Louis Valiquette, Philippe Martin, Simon Labbé

## Abstract

The early diagnosis of severe acute respiratory syndrome coronavirus 2 (SARS-CoV-2) infections is required to identify and isolate contagious patients to prevent further transmission of the coronavirus disease 2019 (COVID-19). In this study, we present a multitarget real-time TaqMan reverse transcription PCR (rRT-PCR) assay for the quantitative detection of SARS-CoV-2 and some of its circulating variants harboring mutations that give SARS-CoV-2 a selective advantage. Seven different primer-probe sets that included probes containing locked nucleic acid (LNA) nucleotides were designed to amplify specific wild-type and mutant sequences in Orf1ab, Envelope (E), Spike (S), and Nucleocapsid (N) genes. Furthermore, a newly developed primer-probe set targeted human β_2_-microglobulin (B2M) as a highly sensitive internal control for RT efficacy. All singleplex and fourplex assays detected ≤ 14 copies/reaction of quantified synthetic RNA transcripts, with a linear amplification range of 9 logarithmic orders. Primer-probe sets for detection of SARS-CoV-2 exhibited no false-positive amplifications with other common respiratory pathogens, including human coronaviruses NL63, 229E, OC43, and HKU-1. Given the emergence of SARS-CoV-2 variants and their rapid spread in some populations, fourplex rRT-PCR assay containing four primer-probe sets represents a reliable approach to detect multiple viral target sequences containing typical mutations of SARS-CoV-2 variants in a single reaction, allowing quicker detection of circulating relevant variants.

## Introduction

The human SARS-CoV-2 virus that causes the respiratory infection COVID-19 started in the Wuhan province of China in late 2019, and then spread around the world (Wu et al. 2020; Zhang and Holmes 2020; Zhu et al. 2020). This novel coronavirus belongs to the subgroup betacoronavirus and the subgenus Sarbecovirus (Chan et al. 2020; Lu et al. 2020a; Zhu et al. 2020). The SARS-CoV-2 genome is constituted of a positive single-stranded RNA with a genome size of nearly ∼29 800 to 29 900 nucleotides in length (Lu et al. 2020a). At the beginning of the outbreak, numerous patients exhibiting an atypical viral pneumonia in Wuhan City were notified to World Health Organization (Wu et al. 2020). Shortly thereafter, infected foreign residents of China and foreign travelers arriving in their countries of residence from international destinations, including visitors from Wuhan, contributed to the spread of the virus through person-to-person contact (Chan et al. 2020; Holshue et al. 2020; Oude Munnink et al. 2020). Subsequently, spreading of the virus occurred easily and rapidly from person-to-person transmission in local communities, resulting in a pandemic spread across the globe.

Although a number of vaccines against SARS-CoV-2 have been developed, they are not available to all people as their production and distribution require efficient logistic and time. Until vaccines become universally available, governments from many countries around the world strongly suggest to their citizens to maintain and sometimes enforce social distancing as they impose lockdown measures, overnight curfew and implement an obligatory quarantine in the case of international travelers flying or driving to different countries. Although rapid identification of SARS-CoV-2-infected patients is critical to trigger off their isolation, the early diagnosis of SARS-CoV-2 remains difficult for the following reasons. First, some SARS-CoV-2-infected patients exhibit no symptoms throughout the course of the infection. Second, other SARS-CoV-2-infected patients with mild symptoms could be confounded with patients who possess other kinds of atypical respiratory tract infections (Guarneri et al. 2021; Lai et al. 2020; Rivett et al. 2020; Yang et al. 2020). In response to the outbreak of COVID-19, numerous national public health agencies across the world have been proactive by setting up testing programs. Most of these agencies have recommended and used a nucleic-acid-based method for early detection of SARS-CoV-2. These nucleotide-based tests rely on the real-time reverse transcription PCR (rRT-PCR) that has been considered the “gold standard” approach for SARS-CoV-2 detection due to its hypersensitivity for correctly identifying the viral RNA genome found in nasopharyngeal swabs and sputum samples collected from infected patients (Corman et al. 2020; Esbin et al. 2020; Lu et al. 2020b).

For daily clinical specimen testing, clinical microbiology laboratories have used different primer-probe sets for detection of SARS-CoV-2 (Corman et al. 2020; Lu et al. 2020b; Nalla et al. 2020). Among them, the CDC N2 and Corman E (also called E Sarbeco) primer-probe sets have been identified to be particularly sensitive (Nalla et al. 2020). A typical singleplex reaction contains one set of primer pairs (forward and reverse) and a TaqMan probe that hybridizes to a specific targeted region of the viral genome. Short sequences within the N and E gene regions are examples of viral templates proved to be detectable with high reproducibility (Corman et al. 2020; Lu et al. 2020b). In the same master mixture, a second set of primer pairs and TaqMan probe containing a distinct fluorophore are added to target a human gene, such as the ribonuclease P gene (RNase P also denoted RP or RPP30) to monitor nucleic acid extraction and amplification (Lu et al. 2020b; Lu et al. 2014).

Since the beginning of the COVID-19 pandemic, several studies have reported genomic sequence variations of SARS-CoV-2 isolates (Leung et al. 2021; Martin et al. 2021; Rondinone et al. 2021; Tegally et al. 2021a; Tegally et al. 2021b). Among these genetic variations, a single nucleotide polymorphism (SNP) found at position 8,782 (in *orf1ab*; C instead of T) of SARS-CoV-2 genomic sequence is a hallmark of the L strain (Tang et al. 2020), whereas the unaltered nucleotide is found at the same position in the S strain. Several mutations have also been found in the coding sequence of the Spike protein (Starr et al. 2020; Yi et al. 2020). One critical mutation in which an adenine has been substituted by a guanine at position 23,403 in the genome of the Wuhan reference strain produces a mutant form of Spike containing D614G substitution (aspartic acid to glycine substitution at amino acid residue 614) (Korber et al. 2020). Another critical mutation in which an adenine has been substituted by a thymine at position 23,063 generates a mutant form of Spike harboring N501Y substitution (asparagine to tyrosine substitution at amino acid residue 501) that is found in the United Kingdom (UK) B.1.1.7 strain as well as other reported variants (Leung et al. 2021). These mutations represent few examples of the ability of SARS-CoV-2 to rapidly evolve since the beginning of the outbreak.

In this study, we report the development of novel primer-probe sets for the detection of SARS-CoV-2 variants. We use fluorogenic probes containing locked nucleic acid (LNA) for detection of single nucleotide polymorphisms (SNPs) in the genome of SARS-CoV-2 using the TaqMan-LNA rRT-PCR method. In addition to the new primer-probe sets, we have developed a fourplex rRT-PCR assay in which four sets of primer pairs and fluorogenic probes are contained in a single reaction master mixture. Furthermore, we have created and used a novel set of primers/probe that detects the ubiquitously expressed human β_2_-microglobulin (B2M) transcripts (instead of RNase P) as a positive control for monitoring performance of the whole procedure. These include the presence of RNAs in the collected sample, RNA extraction, reverse transcription efficacy, and real-time amplification. Thus, this multitarget rRT-PCR assay will improve our ability to narrow the differential identification of SARS-CoV-2 isolates.

## Materials and methods

### Primer and probe design

The Wuhan-Hu-1 genome reference sequence of SARS-CoV-2 (Lu et al. 2020a; Wu et al. 2020) as well as several other listed sequences of its genome (available in the repository of the National Center for Biotechnology Information) were aligned using BLASTn software to identify conserved nucleotide regions corresponding only to the SARS-CoV-2 viral genome and not found in other viral sequences. Subsequently, different primer-probe sets targeting distinct regions of the SARS-CoV-2 genome, especially within E, Orf1ab, and S genes, were designed using Primer Express 3.0.1 software. The N LSPQ primer-probe set was designed as described previously (Longtin et al. 2021). After validation of their exclusivity by comparing them to Virus Pathogen Resource (ViPR), Reference Viral DataBase (RVDB) and human GRCh38 genomic DNA/mRNA databases, the indicated TaqMan primer-probe sets (Table 1) were predicted to specifically amplify SARS-CoV-2, exhibiting the absence of non-specific homologies with other respiratory viral pathogens or human-related gene sequences. The only exception was the E Sarbeco primer-probe set that was predicted to hybridize with the E gene sequence of SARS-CoV-1 in addition to the E gene of SARS-CoV-2 (Corman et al. 2020). TaqMan probes were labeled at the 5’ end with either 6-carboxyfluorescein (6-FAM), Texas Red 615 (TEX 615), hexachlorofluorescein (HEX), or Cytiva5 (Cy5) and at the 3’ end with either Iowa Black FQ (IBFQ) or Black Hole Quencher 1 (BHQ1) (Integrated DNA Technologies, Coralville, Iowa).

**Table 1.**
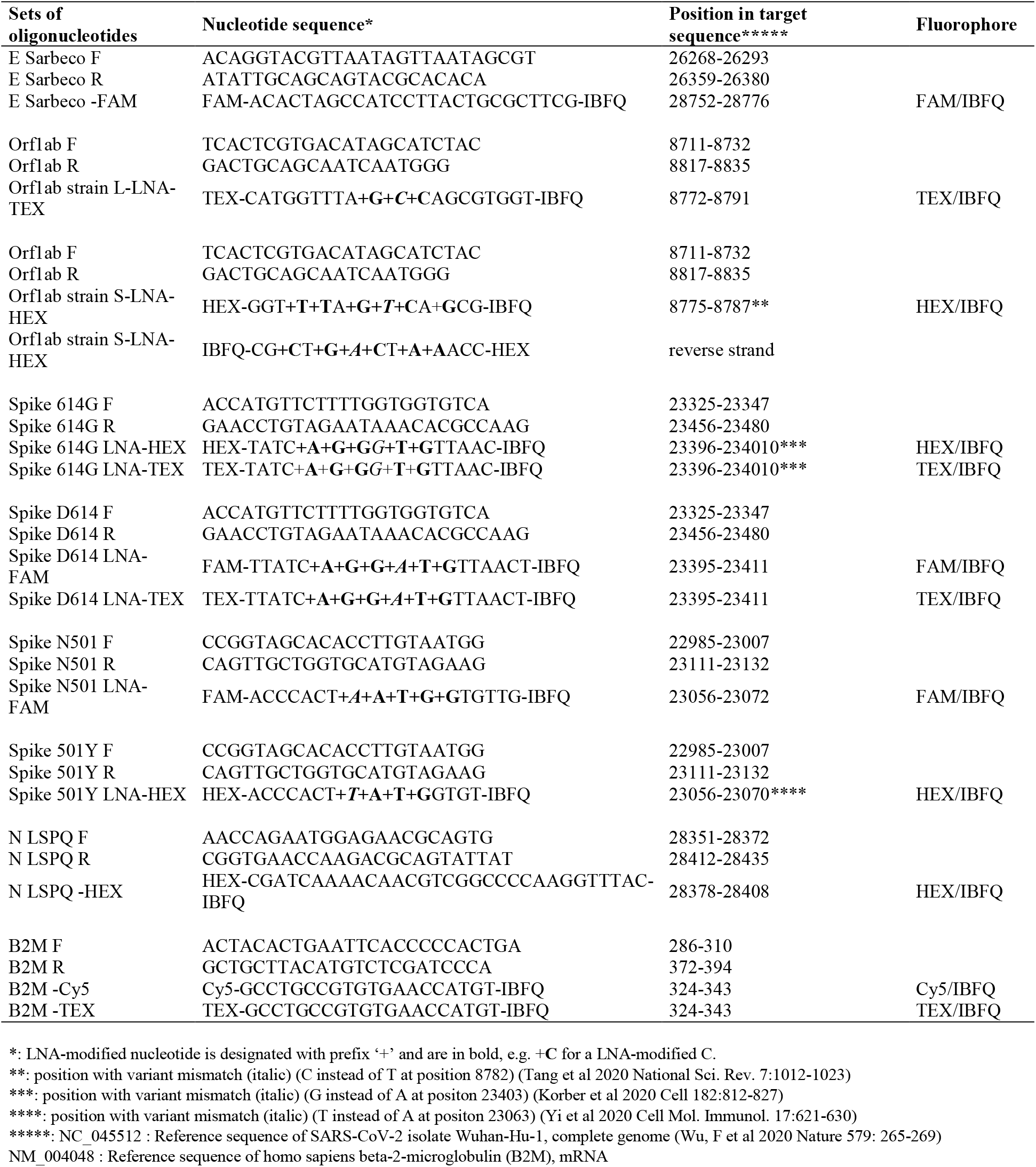
Oligonucleotide primers and fluorogenic probes used in real-time quantitative RT-PCR assays.

In the case of probes containing locked nucleic acid (LNA) residues, they were designed for detection of single nucleotide polymorphisms (SNPs) in the Orf1ab and S genes of SARS-CoV-2. Essentially, we used forward and reverse primers that hybridized with each side of the target sequence and wild-type or mutant TaqMan LNA probes that hybridized at the mutation site, which was located between the forward and reverse primers. All primers and TaqMan probes were designed to avoid self-complementarity and the formation of primer-dimer by-products and hairpins. Furthermore, all primer/probe sets were validated for their compatibility in multiplex rRT-PCR assays.

### Synthesis of RNA transcripts used as standards

gBlocks double-stranded DNA fragments encompassing the amplified region of each targeted SARS-CoV-2 region were synthesized accompanied with a T7 promoter sequence at their 5’ends (Fig. S1). A similar approach was used in the case of the DNA fragment covering the amplified region of the human β-2-microglobulin (B2M) gene. Purified gBlocks products were used for *in vitro* T7-dependent transcription, as described previously (Vijgen et al. 2005). The gBlocks DNA templates were eliminated by digestion with RNase-free DNase I for 15 min at 37°C. The RNA transcripts were purified using a MEGAclear transcription clean-up kit (Invitrogen) and were quantified spectrophotometrically at 260 nm. After RNA levels quantification, measurements of transcripts were converted to the molecule number per µl as described previously (Fronhoffs et al. 2002). To validate calculations of viral RNA copies per reaction, we have established standard curves from 10-fold dilutions of RNA isolated from known number of copies from a prototype viral RNA preparation that was commercially available (Exact Diagnostics, Fort Worth, TX; Vircell, Granada, Spain).

### Real-time quantitative reverse transcription PCR (rRT-PCR)

Reactions were performed in a 10-μl reaction mixture including 7 µl extracted RNA-containing samples or RNA transcripts used as standards, 2.5 µl Reliance One-Step Multiplex rRT-PCR Supermix (4X concentrated) (BioRad), and 0.5 µl of forward and reverse primers (500 nM) and the indicated TaqMan or TaqMan-LNA probe (250 nM). One-step rRT-PCR amplification and detection were performed in a CFX96 Touch Real-Time PCR System (BioRad) under the following conditions. Reverse transcription of 10 min at 50°C was followed by PCR activation at 95°C for 10 min and 50 cycles of amplification (10 s at 95°C and 30 s at 60°C or for some indicated reactions at 63°C). In singleplex reaction mixtures, each reaction mixture contained a single primer pair and TaqMan or TaqMan-LNA probe. In the case of fourplex reactions, they contained all four sets of primers (500 nM per set) and probes (250 nM per probe) within a single reaction master mixture. Fluorescence measurements were monitored after each amplification round and the threshold cycle (C_t_) value for each sample was calculated by assessing the point at which fluorescence crossed the threshold line, exhibiting an increase in fluorescence above the calculated background levels. The result was considered valid if two or more of the target-specific fluorescent signal showed the C_t_ value ≤ 37 cycles and all positive and negative control reactions gave a successful and no amplification, respectively.

### BioFire FilmArray assays

The collection of respiratory viral and bacterial pathogens was obtained from ZeptoMetrix (cat #NATRVP-IDI). The collection is called NATtrol Respiratory Verification Panel (NATRVP) and contains 20 vials x 0.6 ml, each containing viral and bacterial targets listed in Table 4. A master mixture that was constituted of an aliquot of each pathogen of NATRVP was injected into the BioFire Respiratory Panel 2.1 (RP2.1) Pouch in accordance with the manufacturer’s instructions for analyze using the BioFire FilmArray 2.0 System (BioFire Diagnostics) as described previously (Poritz et al. 2011). The loaded RP2.1 Pouch inserted into the FilmArray instrument contained all necessary reagents for automated nucleic acid extraction, reverse transcription, and two consecutive multiplex PCR amplification runs. Furthermore, the FilmArray instrument has the potential to undergo gene target melt curve analysis with each target in a valid run reported as detected or not detected.

## Results

### Limits of detection (LoD) with SARS-CoV-2 RNA transcripts in singleplex reactions

gBlocks DNA templates containing a 5’ T7 RNA polymerase promoter sequence were used to produce viral transcripts that corresponded to specific SARS-CoV-2 genomic regions (Fig. S1). These RNA molecules were used as standards for the generation of standard curves to determine the limit of detection (LoD) of each primer-probe set. Oligonucleotide primer pairs and fluorogenic probes are listed in Table 1. In the case of the E Sarbeco probe and primers, their nucleotide sequences were identical to those described previously (Corman et al. 2020). The other primer pairs and fluorogenic probes were designed from the SARS-CoV-2 complete genome (Lu et al. 2020a; Wu et al. 2020). In the cases of SARS-CoV-2 L and S strains, specific TaqMan locked nucleic acid (LNA) probes were designed to discriminate between the single nucleotide polymorphism (SNP) found at position 8,782 (in *orf1ab*; C instead of T) of SARS-CoV-2 genomic sequence as described previously (Tang et al. 2020). Similarly, a specific TaqMan LNA probe was synthesized to detect a guanine instead of an adenine at position 23,403 in the Wuhan reference strain (Lu et al. 2020a; Wu et al. 2020), allowing the identification of the Spike variant harboring a glycine residue instead of an aspartic acid residue at position 614 (Zhang et al. 2020). An additional mutation found in the UK strain B.1.1.7 was of interest to probe with a specific TaqMan LNA oligonucleotide to discriminate between the SNP found at position 23,063 (in *Spike*; thymine instead of adenine) where a tyrosine residue instead of an asparagine residue at position 501 was found (Kemp et al. 2021). The sensitivity of each set of primer pair and TaqMan probe was first evaluated in singleplex reactions. Tenfold serial dilutions of viral transcript ranging from 1.3 to 1.4 × 10^9^ copies per reaction mixture were tested in triplicate by real-time quantitative RT-PCR (rRT-PCR) assays. Results were analyzed in terms of the C_t_ value that was defined as the threshold cycle in which a target viral sequence was first detected. Samples with C_t_ values ≤ 37 were considered to be positive in comparison with background cross-reactivity of the primers and probes in non-template control (NTC) reactions. The highest dilution of transcript that gave a significative C_t_ value was defined as the limit of detection (LoD) for a targeted RNA transcript. Results showed that LoD values ranged from 1.3 to 13 or 14 RNA transcript copies / reaction (Table 2). Linear regression curves were achieved over a 9-log dynamic range, from 1.3 to 1.3 × 10^9^ copies or 1.4 to 1.4 × 10^9^ copies per reaction for all nine probes, with calculated efficient values of 83.3% to 95.1% (Fig. 1).

**Table 2.** Limits of detection of SARS-CoV-2-specific primers and probes using synthetic RNA transcripts in singleplex rRT-PCR assays.

| Lowest numbers of copies/reaction | Orflab L LNA-TEX |  |  | Orflab S LNA-HEX |  |  |
| --- | --- | --- | --- | --- | --- | --- |
|  | Test 1 (Ct) | Test 2 (Ct) | Test 3 (Ct) | Test 1 (Ct) | Test 2 (Ct) | Test 3 (Ct) |
| 1.4 | 34.83 | 34.78 | 34.81 | 35.22 | 35.67 | 35.37 |
| 14 | 30.73 | 30.55 | 30.61 | 32.64 | 32.86 | 32.71 |
| 140 | 27.11 | 27.07 | 27.02 | 29.46 | 29.27 | 29.35 |
| 1.4E+03 | 23.85 | 23.81 | 23.78 | 25.64 | 25.63 | 25.55 |
| 1.4E+04 | 20.04 | 19.96 | 19.91 | 21.97 | 21.91 | 21.88 |
| 1.4E+05 | 16.55 | 16.51 | 16.47 | 18.54 | 18.55 | 18.47 |
| 1.4E+06 | 12.72 | 12.69 | 12.65 | 15.60 | 15.05 | 15.27 |
| 1.4E+07 | 10.03 | 10.00 | 9.92 | 11.22 | 11.19 | 11.11 |
| 1.4E+08 | 7.03 | 6.52 | 6.72 | 7.76 | 8.01 | 7.91 |
| 1.4E+09 | 3.74 | 3.60 | 3.68 | 4.79 | 5.10 | 4.87 |
| NTC | Neg | Neg | Neg | Neg | Neg | Neg |

| Lowest numbers of copies/reaction | Spike 614G LNA-HEX or TEX |  |  | Spike D614 LNA-FAM or TEX |  |  |
| --- | --- | --- | --- | --- | --- | --- |
|  | Test 1 (Ct) | Test 2 (Ct) | Test 3 (Ct) | Test 1 (Ct) | Test 2 (Ct) | Test 3 (Ct) |
| 1.3 | 34.84 | 34.39 | 34.61 | 36.60 | 36.71 | 36.81 |
| 13 | 32.94 | 31.7 | 31.77 | 33.51 | 34.6 | 33.77 |
| 130 | 29.72 | 29.67 | 29.56 | 30.89 | 30.91 | 30.81 |
| 1.3E+03 | 26.13 | 26.16 | 26.04 | 27.04 | 27.25 | 27.09 |
| 1.3E+04 | 22.61 | 22.64 | 22.55 | 22.52 | 22.56 | 22.47 |
| 1.3E+05 | 18.82 | 18.91 | 18.85 | 18.88 | 18.95 | 18.81 |
| 1.3E+06 | 15.14 | 15.09 | 15.02 | 15.24 | 15.17 | 15.09 |
| 1.3E+07 | 11.48 | 11.53 | 11.41 | 11.47 | 11.46 | 11.34 |
| 1.3E+08 | 7.70 | 7.65 | 7.61 | 7.74 | 7.76 | 7.65 |
| 1.3E+09 | 3.89 | 3.91 | 3.97 | 4.30 | 4.31 | 4.22 |
| NTC | Neg | Neg | Neg | Neg | Neg | Neg |

| Lowest numbers of copies/reaction | Spike N501 LNA-FAM |  |  | Spike 501Y LNA-HEX |  |  |
| --- | --- | --- | --- | --- | --- | --- |
|  | Test 1 (Ct) | Test 2 (Ct) | Test 3 (Ct) | Test 1 (Ct) | Test 2 (Ct) | Test 3 (Ct) |
| 1.4 | 35.55 | 35.76 | 34.51 | nd | nd | nd |
| 14 | 32.09 | 32.28 | 32.51 | 33.98 | 34.83 | 33.60 |
| 140 | 28.99 | 28.73 | 29.38 | 32.42 | 31.71 | 31.58 |
| 1.4E+03 | 25.64 | 25.55 | 25.97 | 27.88 | 27.94 | 28.22 |
| 1.4E+04 | 22.16 | 22.18 | 22.26 | 24.51 | 24.55 | 24.60 |
| 1.4E+05 | 18.58 | 18.54 | 18.54 | 21.01 | 21.04 | 20.95 |
| 1.4E+06 | 14.85 | 14.78 | 14.95 | 17.47 | 17.56 | 17.55 |
| 1.4E+07 | 11.34 | 10.96 | 10.98 | 13.53 | 13.67 | 13.69 |
| 1.4E+08 | 8.40 | 8.45 | 8.35 | 10.25 | 10.4 | 10.29 |
| 1.4E+09 | 4.02 | 3.92 | 4.60 | 6.23 | 6.79 | 6.41 |
| NTC | Neg | Neg | Neg | Neg | Neg | Neg |

**Table 2.** continued
| Lowest numbers of<br>copies/reaction | E Sarbeco-FAM |  |  |
| --- | --- | --- | --- |
|  | Test 1 (Ct) | Test 2 (Ct) | Test 3 (Ct) |
| 1.3 | 35.79 | 36.46 | 35.96 |
| 13 | 32.29 | 32.38 | 32.33 |
| 130 | 28.33 | 28.21 | 28.27 |
| 1.3E+03 | 23.79 | 23.82 | 23.61 |
| 1.3E+04 | 18.53 | 18.52 | 18.63 |
| 1.3E+05 | 14.85 | 14.84 | 14.96 |
| 1.3E+06 | 11.39 | 11.38 | 11.44 |
| 1.3E+07 | 7.62 | 7.61 | 7.81 |
| 1.3E+08 | 5.62 | 5.67 | 5.71 |
| 1.3E+09 | 3.01 | 3.09 | 3.13 |
| NTC | Neg | Neg | Neg |

| Lowest numbers of<br>copies/reaction | N LSPQ-HEX |  |  |
| --- | --- | --- | --- |
|  | Test 1 (Ct) | Test 2 (Ct) | Test 3 (Ct) |
| 1.4 | nd | nd | nd |
| 14 | 36.66 | 36.76 | 36.71 |
| 140 | 32.54 | 32.00 | 32.33 |
| 1.4E+03 | 28.80 | 28.76 | 28.58 |
| 1.4E+04 | 25.30 | 25.36 | 25.26 |
| 1.4E+05 | 21.71 | 21.72 | 21.59 |
| 1.4E+06 | 18.20 | 18.16 | 18.09 |
| 1.4E+07 | 14.65 | 14.64 | 14.51 |
| 1.4E+08 | 11.00 | 11.04 | 11.11 |
| 1.4E+09 | 7.52 | 7.07 | 7.15 |
| NTC | Neg | Neg | Neg |

| Lowest numbers of<br>copies/reaction | B2M-Cy5 or TEX |  |  |
| --- | --- | --- | --- |
|  | Test 1 (Ct) | Test 2 (Ct) | Test 3 (Ct) |
| 1.3 | 32.90 | 32.84 | 32.97 |
| 13 | 30.73 | 30.88 | 30.96 |
| 130 | 27.98 | 27.49 | 27.64 |
| 1.3E+03 | 24.00 | 23.64 | 23.88 |
| 1.3E+04 | 20.43 | 19.77 | 20.05 |
| 1.3E+05 | 16.20 | 16.21 | 16.38 |
| 1.3E+06 | 12.52 | 12.44 | 12.61 |
| 1.3E+07 | 8.64 | 8.69 | 8.72 |
| 1.3E+08 | 7.10 | 6.90 | 6.95 |
| 1.3E+09 | 3.00 | 3.10 | 3.19 |
| NTC | Neg | Neg | Neg |

**Fig. 1.**
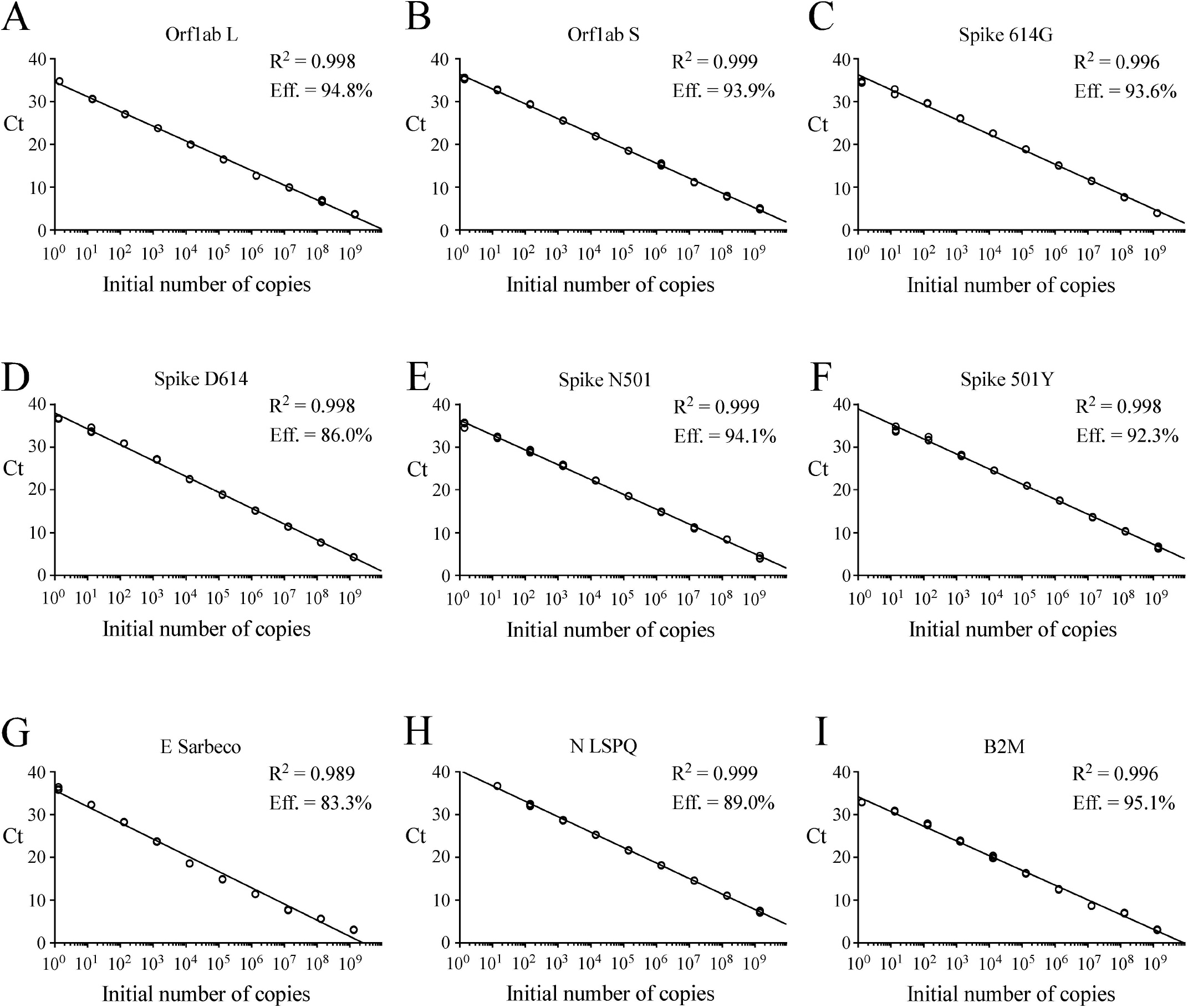
Standard curves to determine synthetic RNA copy number. *A – I*. Tenfold serial dilutions ranging from 10^0^ to 10^9^ copies of Orf1ab L. Orf1ab S. Spike 614G. Spike D614. Spike N501. Spike 501Y. E Sarbeco. N LSPQ. and B2M synthetic RNA transcripts were analyzed by rRT-PCR assays. Each representative graph was generated by plotting the C_t_ values (y-axis) and the log of initial number of copies of synthetic transcripts (x-axis). Calculated linear correlation coefficients (R^2^) and percentage of amplification efficiencies are indicated for each primer-probe set. The graphs represent quantification of the results of three independent experiments.

### Limits of detection (LoD) in the case of fourplex rRT-PCR assays

Following analytical sensitivity of the primers and probes in singleplex reactions, we sought to develop a fourplex rRT-PCR assay that contained four sets of primers and probes within one reaction master mixture. Limits of detection in the indicated fourplex assays were determined to verify whether their sensitivities were comparable to those of singleplex reactions (Tables 2 and 3). Fourplex reactions that were prioritized included TaqMan probes for detection of single nucleotide polymorphisms in the genome of the SARS-CoV-2 strain that had been found in patient samples (Korber et al. 2020; Leung et al. 2021). With the goal of identifying distinct SARS-CoV-2 virus variants, different combinations of primer-probe sets were mixed within one reaction master mixture to specifically detect viral RNA genes that have undergone genetic variations. As an example, a fourplex master mixture contained all four sets of primers and probes to determine whether a SARS-CoV-2 isolate possessed viral RNA that encoded a Spike variant harboring either an Asp^614^ or a Gly^614^ (also denoted D614G mutation) (Korber et al. 2020). Similarly, additional fourplex master mixtures contained sets of primers and probes to determine whether a SARS-CoV-2 isolate belongs to the L or S strain (Tang et al. 2020). Other examples of fourplex reactions included primer-probe sets to identify the SNP found in *Spike gene* at position 23,063 (thymine instead of adenine) that results in an amino acid substitution N501Y that triggers a stronger interaction of the Spike receptor-binding domain with the human cell-surface receptor angiotensin-converting enzyme 2 (ACE2) (Leung et al. 2021; Martin et al. 2021). This multiplex approach could facilitate tracking of SARS-CoV-2 variants that carry mutations. Serial 10-fold dilutions of SARS-CoV-2 transcripts were produced as described above and were tested using different combinations of sets of primer pairs and fluorogenic probes in one reaction mixture as described in Table 3. Linear amplification was performed over a 9-log dynamic range, from 1.4 to 1.4 × 10^9^ copies per tetraplex reaction for all combined probes (Table 3). Results showed linear regression curves that exhibited calculated efficient values of 85.1% to 110.4% (Fig. 2).

**Table 3.** Sensitivities of SARS-CoV-2-specific primers and probes in fourplex rRT-PCR assays.

| Fourplex 1 |  |  |  |  |  |  |  |  |  |  |  |  |
| --- | --- | --- | --- | --- | --- | --- | --- | --- | --- | --- | --- | --- |
| Probes in the fourplex | E Sarbeco-FAM |  |  | Orf1ab L LNA-TEX |  |  | N LSPQ-HEX |  |  | B2M-Cy5 |  |  |
| Lowest numbers of copies/reaction | Test 1 (Ct) | Test 2 (Ct) | Test 3 (Ct) | Test 1 (Ct) | Test 2 (Ct) | Test 3 (Ct) | Test 1 (Ct) | Test 2 (Ct) | Test 3 (Ct) | Test 1 (Ct) | Test 2 (Ct) | Test 3 (Ct) |
| 1.4 | 35.25 | 35.78 | 35.47 | 32.95 | 32.25 | 32.41 | nd | nd | nd | 33.97 | 34.05 | 33.85 |
| 14 | 33.53 | 33.70 | 33.61 | 29.48 | 29.71 | 29.56 | 37.03 | 36.39 | 36.68 | 30.12 | 30.31 | 30.04 |
| 140 | 30.97 | 30.70 | 30.52 | 27.33 | 27.81 | 27.52 | 35.55 | 33.69 | 34.11 | 26.64 | 27.01 | 26.55 |
| 1.4E+03 | 27.52 | 27.34 | 27.42 | 24.86 | 24.92 | 24.79 | 30.90 | 30.66 | 30.77 | 23.48 | 23.50 | 23.31 |
| 1.4E+04 | 23.93 | 23.97 | 23.88 | 21.89 | 21.75 | 21.78 | 27.28 | 27.30 | 27.42 | 19.85 | 19.97 | 19.71 |
| 1.4E+05 | 20.61 | 20.68 | 20.59 | 18.35 | 18.26 | 18.21 | 23.81 | 23.82 | 23.65 | 16.67 | 16.89 | 16.61 |
| 1.4E+06 | 17.17 | 17.11 | 17.08 | 14.91 | 14.86 | 14.77 | 20.27 | 20.24 | 20.14 | 12.55 | 12.76 | 12.52 |
| 1.4E+07 | 13.62 | 13.66 | 13.55 | 11.30 | 11.21 | 11.17 | 16.87 | 16.89 | 16.73 | 9.52 | 9.63 | 9.46 |
| 1.4E+08 | 10.09 | 10.01 | 9.94 | 7.70 | 7.58 | 7.63 | 13.69 | 13.58 | 13.51 | 6.07 | 6.10 | 6.01 |
| 1.4E+09 | 6.86 | 6.70 | 6.66 | 4.12 | 3.79 | 3.86 | 10.17 | 10.21 | 10.07 | 3.19 | 3.14 | 3.08 |
| NTC | Neg | Neg | Neg | Neg | Neg | Neg | Neg | Neg | Neg | Neg | Neg | Neg |

| Fourplex 2 |  |  |  |  |  |  |  |  |  |  |  |  |
| --- | --- | --- | --- | --- | --- | --- | --- | --- | --- | --- | --- | --- |
| Probes in the fourplex | E Sarbeco-FAM |  |  | Orf1ab L LNA-TEX |  |  | Spike 614G LNA-HEX |  |  | B2M-Cy5 |  |  |
| Lowest numbers of copies/reaction | Test 1 (Ct) | Test 2 (Ct) | Test 3 (Ct) | Test 1 (Ct) | Test 2 (Ct) | Test 3 (Ct) | Test 1 (Ct) | Test 2 (Ct) | Test 3 (Ct) | Test 1 (Ct) | Test 2 (Ct) | Test 3 (Ct) |
| 1.4 | 35.58 | 35.46 | 35.51 | 32.72 | 32.61 | 32.66 | 34.92 | 34.75 | 34.81 | 34.44 | 34.36 | 34.31 |
| 14 | 32.67 | 32.85 | 32.72 | 30.20 | 30.32 | 30.24 | 32.57 | 32.83 | 32.66 | 31.68 | 31.40 | 31.48 |
| 140 | 29.67 | 29.55 | 29.61 | 27.49 | 27.42 | 27.36 | 29.56 | 29.77 | 29.61 | 29.39 | 29.43 | 29.36 |
| 1.4E+03 | 25.83 | 25.92 | 25.72 | 25.86 | 25.84 | 25.72 | 26.47 | 26.8 | 26.62 | 26.91 | 26.82 | 26.78 |
| 1.4E+04 | 22.53 | 22.70 | 22.61 | 22.84 | 23.05 | 22.75 | 21.85 | 22.20 | 21.94 | 23.60 | 23.61 | 23.52 |
| 1.4E+05 | 18.10 | 18.90 | 18.44 | 19.71 | 20.10 | 19.83 | 18.40 | 18.22 | 18.03 | 19.68 | 18.52 | 18.43 |
| 1.4E+06 | 14.96 | 14.93 | 14.84 | 15.32 | 15.60 | 15.28 | 14.55 | 14.60 | 14.42 | 15.32 | 15.45 | 15.25 |
| 1.4E+07 | 11.25 | 11.26 | 11.13 | 11.92 | 11.8 | 11.85 | 10.72 | 10.62 | 10.58 | 11.64 | 11.62 | 11.51 |
| 1.4E+08 | 7.99 | 7.54 | 7.41 | 8.23 | 8.20 | 8.16 | 7.74 | 7.80 | 7.69 | 7.90 | 7.91 | 7.86 |
| 1.4E+09 | 5.10 | 5.25 | 5.21 | 5.54 | 5.56 | 5.45 | 4.20 | 4.01 | 4.07 | 4.95 | 4.92 | 4.87 |
| NTC | Neg | Neg | Neg | Neg | Neg | Neg | Neg | Neg | Neg | Neg | Neg | Neg |

| Fourplex 3 |  |  |  |  |  |  |  |  |  |  |  |  |
| --- | --- | --- | --- | --- | --- | --- | --- | --- | --- | --- | --- | --- |
| Probes in the fourplex | E Sarbeco-FAM |  |  | Orf1ab S LNA-HEX |  |  | Spike D614 LNA-TEX |  |  | B2M-Cy5 |  |  |
| Lowest numbers of copies/reaction | Test 1 (Ct) | Test 2 (Ct) | Test 3 (Ct) | Test 1 (Ct) | Test 2 (Ct) | Test 3 (Ct) | Test 1 (Ct) | Test 2 (Ct) | Test 3 (Ct) | Test 1 (Ct) | Test 2 (Ct) | Test 3 (Ct) |
| 1.4 | 36.96 | 36.89 | 36.83 | nd | nd | nd | nd | nd | nd | 32.67 | 32.85 | 32.51 |
| 14 | 32.14 | 32.61 | 32.28 | 32.71 | 32.92 | 32.84 | 33.77 | 34.60 | 34.11 | 29.31 | 29.41 | 29.27 |
| 140 | 28.78 | 28.91 | 28.72 | 30.46 | 30.38 | 30.27 | 31.08 | 31.14 | 31.01 | 26.47 | 26.49 | 26.31 |
| 1.4E+03 | 26.11 | 26.70 | 26.34 | 27.92 | 28.15 | 28.02 | 28.71 | 29.22 | 28.94 | 24.37 | 24.10 | 24.04 |
| 1.4E+04 | 21.77 | 21.73 | 21.59 | 25.98 | 26.51 | 26.12 | 24.37 | 24.23 | 24.18 | 19.33 | 19.32 | 19.22 |
| 1.4E+05 | 17.88 | 17.59 | 17.48 | 21.69 | 21.65 | 21.57 | 20.17 | 20.01 | 19.92 | 16.48 | 16.55 | 16.39 |
| 1.4E+06 | 13.77 | 13.83 | 13.65 | 17.64 | 17.61 | 17.49 | 15.91 | 15.93 | 15.82 | 13.85 | 12.54 | 12.45 |
| 1.4E+07 | 11.10 | 10.88 | 10.77 | 13.26 | 13.45 | 13.31 | 13.60 | 13.34 | 13.41 | 10.85 | 10.90 | 10.70 |
| 1.4E+08 | 7.92 | 7.89 | 7.81 | 10.63 | 10.41 | 10.52 | 10.83 | 10.70 | 10.68 | 7.12 | 7.07 | 7.01 |
| 1.4E+09 | 5.20 | 5.05 | 5.09 | 7.20 | 7.08 | 7.13 | 7.31 | 7.34 | 7.23 | 4.51 | 4.56 | 4.39 |
| NTC | Neg | Neg | Neg | Neg | Neg | Neg | Neg | Neg | Neg | Neg | Neg | Neg |

Table 3. continued
| Fourplex 4 |  |  |  |  |  |  |  |  |  |  |  |  |
| --- | --- | --- | --- | --- | --- | --- | --- | --- | --- | --- | --- | --- |
| Probes in the fourplex | E Sarbeco-FAM |  |  | Orflab L LNA-TEX |  |  | Orflab S LNA-HEX |  |  | B2M-Cy5 |  |  |
| Lowest numbers of copies/reaction | Test 1 (Ct) | Test 2 (Ct) | Test 3 (Ct) | Test 1 (Ct) | Test 2 (Ct) | Test 3 (Ct) | Test 1 (Ct) | Test 2 (Ct) | Test 3 (Ct) | Test 1 (Ct) | Test 2 (Ct) | Test 3 (Ct) |
| 1.4 | 34.75 | 35.88 | 35.70 | 33.33 | 34.52 | 34.47 | 34.56 | 33.83 | 33.97 | 34.40 | 34.44 | 34.57 |
| 14 | 32.44 | 32.35 | 32.29 | 30.53 | 32.14 | 32.26 | 32.76 | 32.07 | 32.21 | 31.63 | 31.65 | 31.77 |
| 140 | 29.88 | 29.82 | 29.69 | 27.73 | 27.76 | 27.58 | 29.84 | 29.93 | 29.76 | 29.11 | 29.01 | 29.22 |
| 1.4E+03 | 25.17 | 25.13 | 25.01 | 24.97 | 25.41 | 25.12 | 27.48 | 27.50 | 27.39 | 26.52 | 26.54 | 26.62 |
| 1.4E+04 | 22.11 | 22.26 | 22.02 | 22.28 | 22.33 | 22.19 | 23.75 | 23.77 | 23.64 | 22.37 | 22.33 | 22.47 |
| 1.4E+05 | 18.87 | 18.54 | 18.42 | 19.28 | 19.72 | 19.46 | 20.72 | 21.07 | 20.87 | 18.69 | 19.13 | 19.22 |
| 1.4E+06 | 15.92 | 15.70 | 15.61 | 16.35 | 16.52 | 16.29 | 17.66 | 17.52 | 17.33 | 15.37 | 15.35 | 15.42 |
| 1.4E+07 | 11.07 | 10.97 | 10.88 | 13.45 | 13.60 | 13.32 | 14.56 | 14.48 | 14.39 | 13.07 | 12.99 | 13.21 |
| 1.4E+08 | 7.88 | 7.94 | 7.74 | 8.32 | 8.34 | 8.19 | 8.96 | 8.99 | 8.84 | 9.20 | 9.22 | 9.37 |
| 1.4E+09 | 4.91 | 5.05 | 4.85 | 6.44 | 6.46 | 6.32 | 6.42 | 6.45 | 6.35 | 6.86 | 5.53 | 6.03 |
| NTC | Neg | Neg | Neg | Neg | Neg | Neg | Neg | Neg | Neg | Neg | Neg | Neg |

| Fourplex 5 |  |  |  |  |  |  |  |  |  |  |  |  |
| --- | --- | --- | --- | --- | --- | --- | --- | --- | --- | --- | --- | --- |
| Probes in the fourplex | N LSPQ-HEX |  |  | Spike D614 LNA-FAM |  |  | Spike 614G LNA-TEX |  |  | B2M-Cy5 |  |  |
| Lowest numbers of copies/reaction | Test 1 (Ct) | Test 2 (Ct) | Test 3 (Ct) | Test 1 (Ct) | Test 2 (Ct) | Test 3 (Ct) | Test 1 (Ct) | Test 2 (Ct) | Test 3 (Ct) | Test 1 (Ct) | Test 2 (Ct) | Test 3 (Ct) |
| 1.4 | nd | nd | nd | nd | nd | nd | nd | nd | nd | 33.65 | 33.85 | 33.71 |
| 14 | 36.68 | 36.81 | 36.72 | 34.88 | 35.94 | 34.76 | 33.95 | 34.15 | 34.03 | 32.68 | 32.71 | 32.59 |
| 140 | 33.20 | 33.46 | 33.31 | 31.70 | 32.43 | 32.01 | 31.72 | 31.83 | 31.66 | 30.52 | 30.54 | 30.41 |
| 1.4E+03 | 29.40 | 29.65 | 29.52 | 28.99 | 29.16 | 28.89 | 28.45 | 28.76 | 28.37 | 26.68 | 26.71 | 26.52 |
| 1.4E+04 | 24.67 | 24.75 | 24.69 | 25.11 | 25.46 | 25.22 | 24.68 | 24.88 | 24.55 | 23.45 | 23.48 | 23.33 |
| 1.4E+05 | 20.66 | 20.83 | 20.71 | 20.37 | 20.44 | 20.28 | 19.82 | 19.97 | 19.69 | 19.32 | 19.56 | 19.22 |
| 1.4E+06 | 17.15 | 17.17 | 17.05 | 16.70 | 16.84 | 16.62 | 16.46 | 16.7 | 16.39 | 14.10 | 14.22 | 14.02 |
| 1.4E+07 | 14.04 | 14.00 | 13.92 | 12.91 | 12.95 | 12.81 | 12.62 | 12.65 | 12.51 | 11.22 | 11.39 | 11.13 |
| 1.4E+08 | 11.15 | 11.21 | 11.08 | 9.58 | 9.57 | 9.41 | 9.29 | 9.31 | 9.19 | 8.41 | 8.47 | 8.32 |
| 1.4E+09 | 7.91 | 7.98 | 7.84 | 6.21 | 6.25 | 6.11 | 6.10 | 6.14 | 6.02 | 5.02 | 5.04 | 4.94 |
| NTC | Neg | Neg | Neg | Neg | Neg | Neg | Neg | Neg | Neg | Neg | Neg | Neg |

| Fourplex 6 |  |  |  |  |  |  |  |  |  |  |  |  |
| --- | --- | --- | --- | --- | --- | --- | --- | --- | --- | --- | --- | --- |
| Probes in the fourplex | E Sarbeco-FAM |  |  | Spike 501Y LNA-HEX |  |  | Spike 614G LNA-TEX |  |  | B2M-Cy5 |  |  |
| Lowest numbers of copies/reaction | Test 1 (Ct) | Test 2 (Ct) | Test 3 (Ct) | Test 1 (Ct) | Test 2 (Ct) | Test 3 (Ct) | Test 1 (Ct) | Test 2 (Ct) | Test 3 (Ct) | Test 1 (Ct) | Test 2 (Ct) | Test 3 (Ct) |
| 1.4 | nd | nd | 35.64 | nd | 35.54 | 35.69 | nd | nd | nd | 33.31 | 33.33 | 33.29 |
| 14 | 33.71 | 33.61 | 33.66 | 32.49 | 31.92 | 32.94 | 33.77 | 34.05 | 33.06 | 30.94 | 30.57 | 30.58 |
| 140 | 30.84 | 30.85 | 30.90 | 29.73 | 29.70 | 29.95 | 31.51 | 31.11 | 31.14 | 28.41 | 28.48 | 28.49 |
| 1.4E+03 | 27.64 | 27.55 | 27.64 | 26.63 | 26.55 | 26.64 | 27.91 | 27.88 | 27.79 | 25.43 | 25.43 | 25.50 |
| 1.4E+04 | 24.02 | 23.95 | 23.95 | 22.88 | 22.88 | 22.87 | 24.25 | 24.21 | 24.27 | 21.64 | 21.86 | 21.60 |
| 1.4E+05 | 20.45 | 20.45 | 20.30 | 19.64 | 19.67 | 19.64 | 20.91 | 20.94 | 20.79 | 18.03 | 17.62 | 17.56 |
| 1.4E+06 | 17.12 | 17.09 | 16.98 | 15.88 | 15.96 | 15.72 | 17.19 | 17.26 | 16.94 | 14.39 | 14.39 | 14.31 |
| 1.4E+07 | 13.60 | 13.59 | 13.38 | 12.21 | 12.06 | 12.09 | 13.29 | 13.11 | 13.18 | 11.46 | 11.56 | 11.40 |
| 1.4E+08 | 9.85 | 9.74 | 9.75 | 9.33 | 9.28 | 9.24 | 10.78 | 10.77 | 10.71 | 6.83 | 6.40 | 6.51 |
| 1.4E+09 | 7.01 | 6.93 | 6.85 | 6.91 | 6.84 | 6.96 | 7.65 | 7.61 | 7.65 | 3.61 | 4.00 | 3.51 |
| NTC | Neg | Neg | Neg | Neg | Neg | Neg | Neg | Neg | Neg | Neg | Neg | Neg |

**Table 3.** continued
| **Fourplex 7** |  |  |  |  |  |  |  |  |  |  |  |  |
| --- | --- | --- | --- | --- | --- | --- | --- | --- | --- | --- | --- | --- |
| **Probes in the fourplex** | **N LSPQ-HEX** |  |  | **Spike N501 LNA-FAM** |  |  | **Spike 614G LNA-TEX** |  |  | **B2M-Cy5** |  |  |
| **Lowest numbers of copies/reaction** | **Test 1 (Ct)** | **Test 2 (Ct)** | **Test 3 (Ct)** | **Test 1 (Ct)** | **Test 2 (Ct)** | **Test 3 (Ct)** | **Test 1 (Ct)** | **Test 2 (Ct)** | **Test 3 (Ct)** | **Test 1 (Ct)** | **Test 2 (Ct)** | **Test 3 (Ct)** |
| 1.4 | nd | nd | nd | 35.02 | nd | nd | nd | 36.2 | nd | 33.42 | 33.52 | 33.92 |
| 14 | 36.29 | 36.89 | 36.71 | 33.77 | 34.20 | 34.92 | 33.46 | 33.97 | 32.80 | 31.46 | 31.39 | 31.32 |
| 140 | 34.27 | 34.36 | 34.45 | 31.18 | 31.51 | 31.28 | 30.72 | 30.76 | 30.69 | 28.77 | 29.03 | 28.76 |
| 1.4E+03 | 31.20 | 31.13 | 31.20 | 27.59 | 27.46 | 27.69 | 27.43 | 27.65 | 27.76 | 25.44 | 25.48 | 25.52 |
| 1.4E+04 | 27.75 | 27.71 | 27.75 | 23.84 | 24.48 | 24.34 | 24.22 | 24.07 | 24.17 | 21.4 | 21.96 | 22.24 |
| 1.4E+05 | 24.02 | 24.12 | 24.00 | 20.65 | 20.55 | 20.61 | 20.29 | 20.52 | 20.27 | 18.23 | 18.17 | 18.21 |
| 1.4E+06 | 20.56 | 20.58 | 20.51 | 16.73 | 16.90 | 16.81 | 16.6 | 16.61 | 16.53 | 14.21 | 14.27 | 14.29 |
| 1.4E+07 | 17.01 | 17.10 | 17.05 | 13.15 | 13.09 | 13.05 | 12.86 | 12.79 | 12.85 | 10.91 | 11.04 | 11.09 |
| 1.4E+08 | 13.69 | 13.66 | 13.74 | 9.64 | 9.24 | 9.61 | 10.31 | 10.26 | 10.37 | 7.46 | 7.40 | 7.53 |
| 1.4E+09 | 10.10 | 10.12 | 10.13 | 6.49 | 6.50 | 6.58 | 7.53 | 7.54 | 7.53 | 4.46 | 4.34 | 4.45 |
| NTC | Neg | Neg | Neg | Neg | Neg | Neg | Neg | Neg | Neg | Neg | Neg | Neg |

**Table 4.**
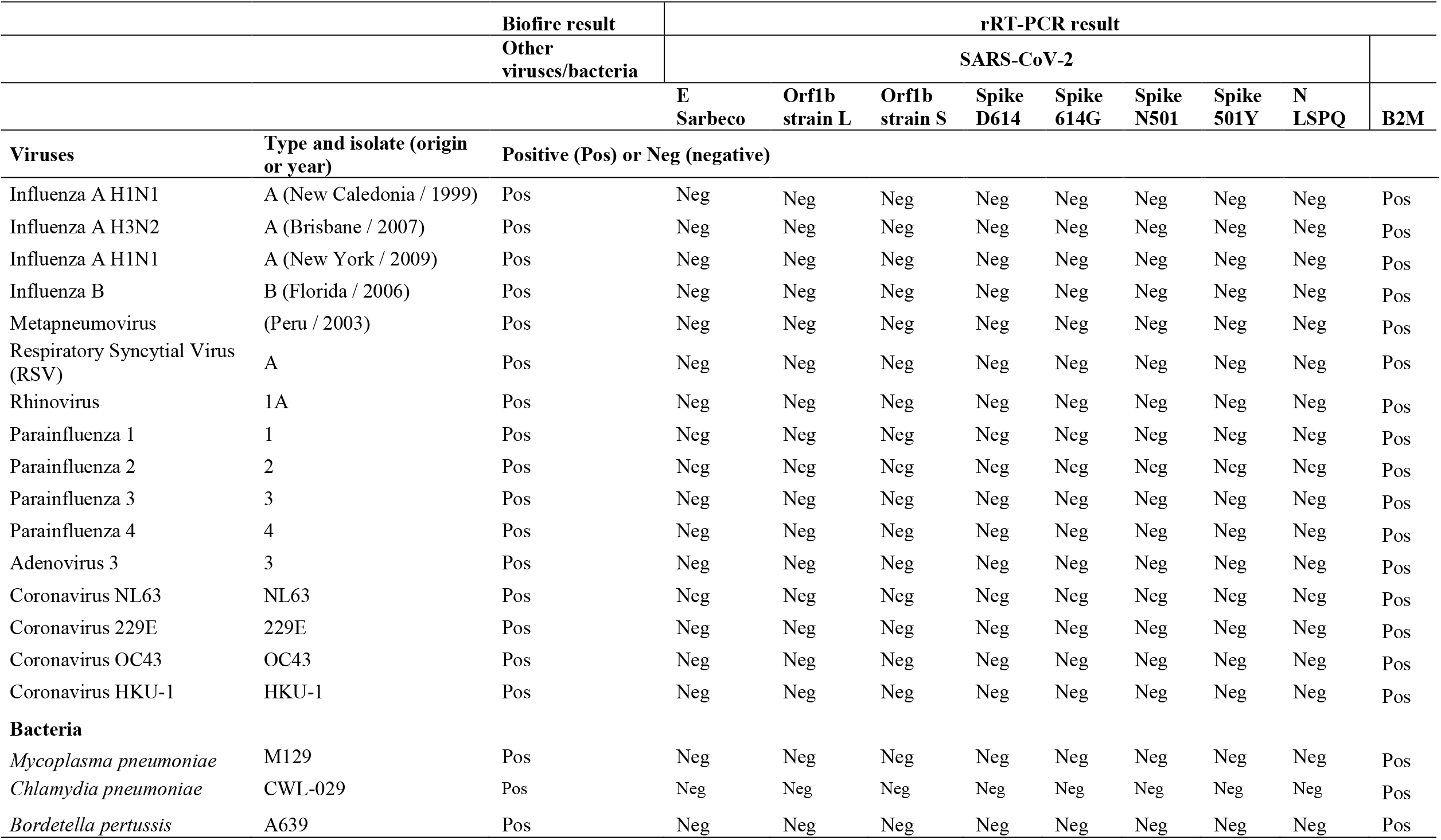
Specificities of fluorogenic SARS-CoV-2 probes with a number of other viruses and bacteria.

**Fig. 2.**
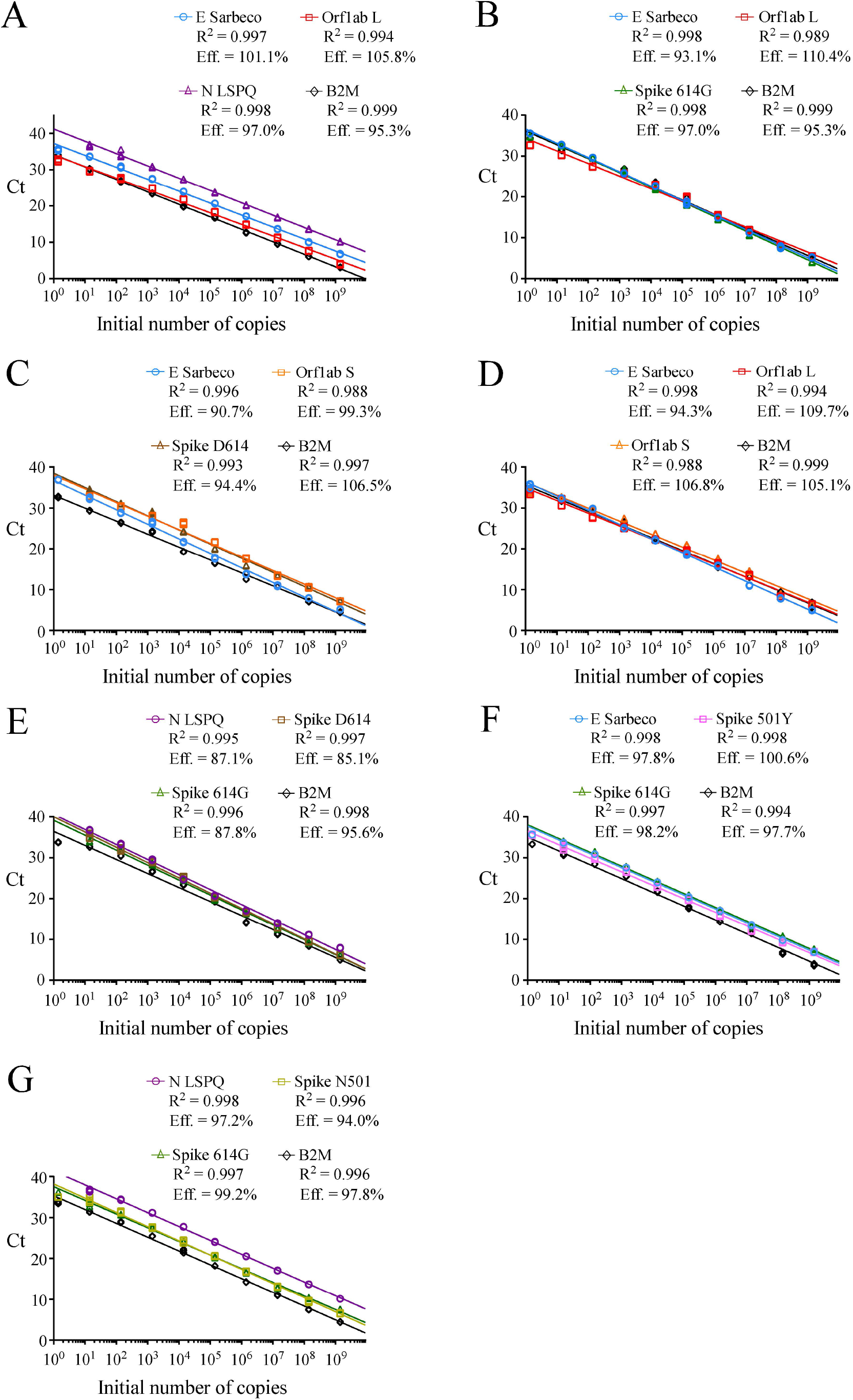
Linear regression curves to assess synthetic RNA copy number in fourplex rRT-PCR assays. *A – G*. For each representative graph. four primer-probe sets within one reaction mixture were used to detect serial 10-fold dilutions of four synthetic RNA transcripts ranging from 10^0^ to 10^9^ copies per reaction. The y and x axes of each graph are the C_t_ values and the log of the input of synthetic RNA transcripts. respectively. For each fourplex reaction. R^2^ indicates calculated linear coefficients and Eff. shows percentage of amplification efficiencies for each primer-probe set that is present with three other sets of primers and probes within one reaction master mixture. Fluorogenic probe color codes are as follows: blue. E Sarbeco; red. Orf1ab L; violet. N LSPQ; black. B2M; green. Spike 614G; orange. Orf1ab S; brown. Spike D614; pink. Spike 501Y; and gold. Spike N501. The graphs represent quantification of the results of three independent experiments.

When we compared the sensitivity measured as the lowest C_t_ value of the TaqMan probes (Orf1ab L, Orf1ab S, Spike 614G, Spike D614, Spike N501, Spike 501Y, E Sarbeco, N LSPQ, and B2M) in singleplex reactions (Table2) and fourplex reactions (Table 3), results showed that sensitivities of these probes between singleplex and fourplex reactions were comparable. In the cases of E Sarbeco, Orf1ab L, and B2M, their limit of detection was generally 1.3 or 1.4 copies of transcript per reaction in both singleplex and fourplex rRT-PCR assays. In the cases of Orf1ab S, Spike D614, Spike 614G, Spike N501, Spike 501Y, and N LSPQ probes, a limit of detection of 13 or 14 copies per reaction was observed with much more consistency in singleplex and fourplex assays. Taken together, all designed primer-probe sets performed comparably in singleplex and fourplex assays that reproducibly detected as few as 1.4 to 14 copies of target sequences per reaction.

### Specificity of primers and probes in fourplex reactions

In the course of designing primer and probe sequences used for detection of SARS-CoV-2, we performed BLAST analyses to verify the absence of significant sequence homologies with other respiratory viruses and human genome sequences to avoid the possibility of false positive results. To test the specificity of the primers and probes that we have used in fourplex rRT-PCR reactions, we performed several assays using aliquots of the entire collection of NATtrol Respiratory Verification Panel (NATRVP) from ZeptoMetrix, which contains purified intact respiratory virus and bacteria particles that have been inactivated to make them non-infectious. These microbial particles were supplied in liquid samples containing a specialized matrix that included human cells to mimic the composition of a true clinical specimen. All SARS-CoV-2 primer and probe sets used in fourplex reactions (see Table 3 for all combinations of four sets of primer and probes per one reaction master mixture that have been tested) showed the absence of nonspecific amplification against the ZeptoMetrix NATRVP preparation that includes 19 respiratory pathogens such as influenza A and B, different types of parainfluenza, respiratory syncytial virus A, and other coronaviruses such as NL63, 229E, OC43, and HKU-1 (Table 4). In contrast, the B2M primer-probe set yielded positive results for the same biological samples due to the presence of human cells that are associated with the ZeptoMetrix NATtrol Respiratory pathogen samples. As positive assays for nucleic acid detection of the 19 respiratory pathogens of the ZeptoMetrix NATRVP, we performed BioFire tests using the BioFire FilmArray 2.0 system (BioFire Diagnostics, Salt Lake City, UT). A master mixture containing all viral and bacterial particles of ZeptoMetrix was injected into the BioFire Respiratory Panel 2.1 Pouch that contained all the necessary reagents for automated nuclei acid extraction, reverse transcription, two stages PCR amplification, and detection of multiple respiratory pathogen targets in a single assay. Results showed that the FilmArray instrument in combination with BioFire Pouch reagents including multiple independent sets of primers allowed multiplex-tandem PCR detection of the microbial panel members listed in Table 4. Therefore, FilmArray runs validated the presence of viral and bacterial nucleic acids in the ZeptoMetrix NATRVP preparation. Taken together, the results showed that fourplex assays are specific for the SARS-CoV-2 virus target and that there is an absence of false-positive signals with other respiratory viral or bacterial pathogens.

### LNA probes allow differential identification of specific SARS-CoV-2 transcripts in fourplex reactions

We tested different groups of differentially labeled four-color sets of probes that allow discrimination of single-nucleotide mutations found in genetic variants of SARS-CoV-2 to validate specific LNA probe-target interaction for SNPs detection in the genome of SARS-CoV-2 (Table 5). Results showed that the Orf1ab L probe containing a “C LNA” at the polymorphic site (position 8782 in SARS-CoV-2 sequence; Table 1) generated a positive fluorogenic signal that was specific for transcripts produced from the new circulating L variant strain, whereas no fluorogenic signal was detected in the presence of RNA obtained from the older S variant strain (Table 5; *e.g*. probes in fourplexes 1 and 2). In the case of the Orf1ab S probe containing a “T LNA” at the polymorphic site (Table 1), this probe exhibited a positive amplification signal in the presence of transcripts produced from the S variant strain and failed to detect transcripts of the L variant strain (Table 5; *e.g*. probes in fourplexes 3 and 4).

**Table 5.**
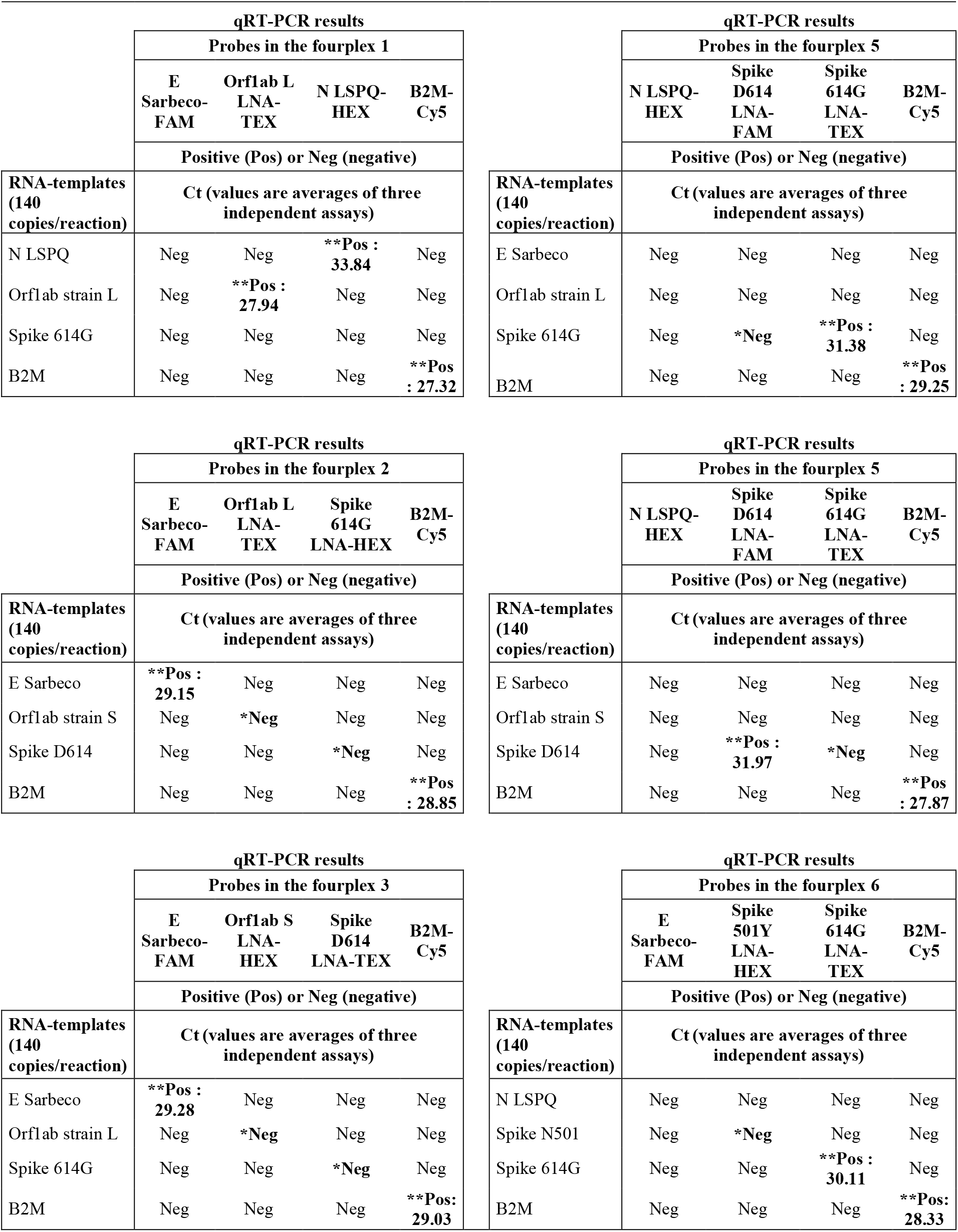
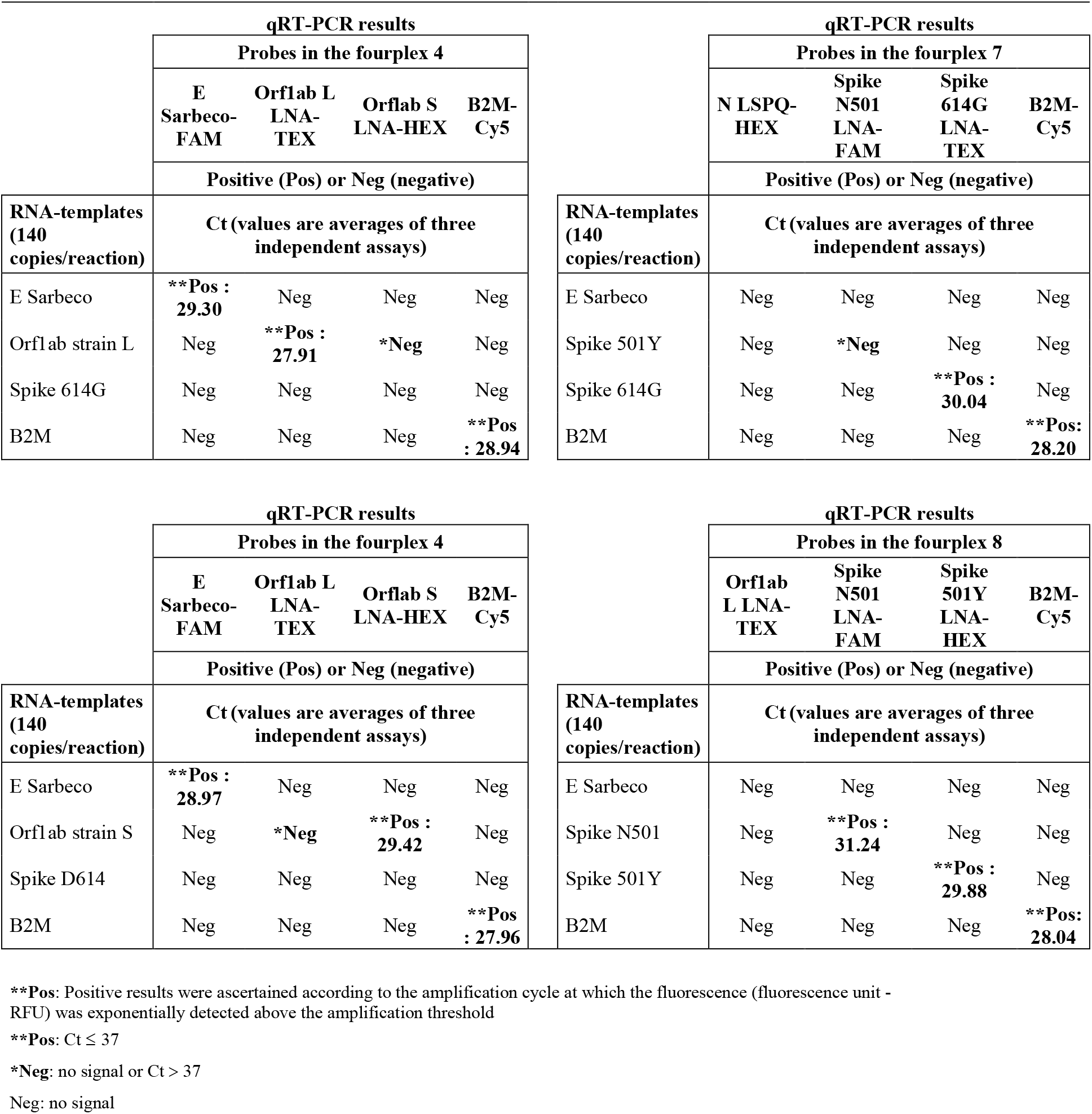
Real-time genotyping with oligonucleotide probes containing locked nucleic acids (LNA) for detection of SARS-CoV-2 variants.

Two oligonucleotide probes containing LNA residues were created for specific hybridization of target sequences containing nucleotide substitutions that confer amino acid substitution in Spike at Asp614 (also denoted D614) and Gly614 (also called 614G) (Table1). The LNA probe for the detection of the Wuhan reference D614 strain carried an “A LNA” at the SNP site (position 23,403 in the viral genome), whereas the fluorogenic probe specific for the detection of the circulating Spike variant (614G) contained a “G LNA” at this position (Table 1). In the case of the Spike D614 LNA probe, results showed that it was specific for the detection of transcripts from the Spike D614 gene and did not generate amplification signal in the presence of transcripts of the Spike 614G variant gene (Table 5; *e.g*. probes in fourplexes 3 and 5). In contrast, results showed that the Spike 614G LNA probe exhibited high specificity for detection of transcripts from the Spike 614G gene but failed to amplify transcripts from the Spike D614 coding sequence (Table 5; *e.g*. probes in fourplex 5). Similarly, experiments by fourplex rRT-PCR assays verified that the LNA probe for the detection of Spike 501Y transcripts (nucleotide change A23,063T) (Tables 1 and 2) failed to detect the Spike N501 transcripts (Table 5, *e.g*. probes in fourplex 6). Consistently, results showed that the LNA probe for detection of the Spike N501 transcripts (nucleotide A at position 23,063) (Tables 1 and 2) did not detect the Spike 501Y transcripts (Table 5, *e.g*. probes in fourplex 7). In contrast, results showed that the Spike N501 and Spike 501Y LNA probes exhibited high specificity for detection of transcripts from the Spike N501 and Spike 501Y coding sequences, respectively (Table 5, *e.g*. probes in fourplex 8). Taken together, the results showed that the oligonucleotide probes containing LNA residues can be used for differential identification of SARS-CoV-2 viral templates in which mutations have occurred.

## Discussion

The ongoing COVID-19 pandemic characterized by successive waves of infection is a driving force for optimization and constant development of SARS-CoV-2 testing assays (Esbin et al. 2020; Graham et al. 2021). Here, we report the development of a fourplex TaqMan-LNA rRT-PCR assay that included per single reaction: four sets of primer pairs and four distinct TaqMan probes labelled with different fluorophores. Results showed that a tetraplex reaction in a single master mixture reached a detection limit of 14 viral transcript copies per reaction (Table 3). The amplification efficiencies of tetraplex reactions (values of 85.1% to 110.4%) were equivalent to those in the singleplex reactions (values of 83.3% and 95.1%) in which a single viral primer-probe set was used within one reaction master mixture (Table 2). The primer-probe set targeting a single nucleotide substitution in the Orf1ab L transcript was more sensitive than primer-probe sets targeting single nucleotide substitutions in the Orf1ab S, Spike 614G, Spike D614, Spike N501, and Spike 501Y transcripts. This set could detect transcript at levels as low as 1.4 copies per reaction in singleplex and fourplex rRT-PCR assays (Tables 2 and 3). In the cases of primer-probe sets targeting single nucleotide substitutions in the Orf1ab S, Spike 614G, Spike D614, Spike N501, and Spike 501Y transcripts, a detection limit of 14 transcript copies was reached when they were used in fourplex assays on three independent experiments (Table 3). In the case of the primer-probe set targeting the E gene that is denoted E Sarbeco (Corman et al. 2020), it was more sensitive than the N LSPQ probe targeting the N gene (Tables 2 and 3). The E Sarbeco primer-probe set could detect transcripts levels as low as 1.3 to 1.4 copies per reaction, whereas 14 transcript copies per reaction was reached with the N LSPQ primer-probe set under our experimental conditions.

During the course of studies, we have found that the human β_2_-microglobulin (B2M) gene encoding an ubiquitously expressed protein could be a positive control of choice for monitoring efficiency of RNA extraction from swab samples and subsequent reverse transcription reaction reliability. To create the B2M primer-probe set and ensure that its resulting product originates from RNA/complementary DNA (cDNA) amplification and not from genomic DNA amplification, we designed the reverse primer on exon2-exon3 junction of the B2M gene. Because this exon2-exon3 boundary sequence is only present in spliced B2M mRNA and not in genomic DNA, this approach eliminated the possibility that the B2M rRT-PCR signal resulted from an amplification of genomic DNA. Results showed that the B2M primer-probe set was highly sensitive when tested in singleplex assays, exhibiting an efficiency of 95.1% (r^2^ = 0.996) and a detection limit of 1.3 transcript copies per reaction (Fig. 1 and Table 2). When the B2M primer-probe set was used in fourplex assays, it remained highly sensitive with high levels of rRT-PCR efficiency (95.5% to 106.5%) and a similar detection limit (1.4 transcript copies per reaction) as compared to that observed in the case of singleplex assays (Figs 1 and 2) (Tables 2 and 3). The vast majority of COVID-19 tests are currently targeting the human RNase P gene (denoted RPP30 or h-RP) as a positive control to monitor RNA extraction and its quality to be reverse transcribed for its subsequent PCR amplification (Lu et al. 2020b; Lu et al. 2014). However, problems with the use of one CDC-recommended h-RP primer-probe set have been reported previously (Dekker et al. 2020). One issue is that the reverse primer hybridizes in the same exon as the forward primer and the TaqMan probe, therefore allowing amplification from genomic DNA instead of exclusively from spliced mRNA (Dekker et al. 2020). In that case, a positive result with this specific primer and probe set fails to validate the quality of the clinical sample specimen with respect to its RNA content.

The use of multiplex rRT-PCR assays allowed simultaneous targeting of different regions of the SARS-CoV-2 genome in a single master mixture. Among the targeted regions, we often included the E Sarbeco probe due to its reported successful use and high sensitivity (Corman et al. 2020; Nalla et al. 2020). In this manner, C_t_ values of the E Sarbeco probe in fourplex assays served as a point of reference to assess the significance of C_t_ values of the other tested probes. Interestingly, we have observed that utilization of multiplex SARS-CoV-2 primer/probe sets with distinct fluorophores shed light on which regions of the virus genome are more sensitive for a rRT-PCR-based SARS-CoV-2-specific identification assay. Another advantage of using a multitarget rRT-PCR method is the fact that it reduces the amount of non-enzymatic reagents, enzyme mixtures and plasticware required to analyze all the samples on a daily basis.

RNA viruses exhibit high mutation rates that may result in self beneficial properties by increasing their ability to infect host cells (Cuevas et al. 2015; Geller et al. 2016; Lauring et al. 2013). In the case of SARS-CoV-2, several mutations have been identified since the start of the pandemic (Fontanet et al. 2021; Mascola et al. 2021; Tegally et al. 2021b). For instance, two linked SNPs in the genome of SARS-CoV-2 have defined two major lineage of the virus that are designated S and L strains (Tang et al. 2020). Another example consists of nucleotide mutations found in the viral sequence of the gene encoding Spike that result in increased virus infectivity, especially nucleotide changes resulting in amino acid changes at Asp614 (D614G) and Asn501 (N501Y) (Korber et al. 2020; Leung et al. 2021). Here, we have used forward and reverse primers that hybridize to each side of hot spot mutational sites found in Orf1ab (nucleotide change T8782C, generating lineage S versus L), and at different locations in Spike such as nucleotide change A23403G (resulting in Spike variation D614G) and nucleotide change A23063T (giving rise Spike variation N501Y). These pairs of primers were used in combination with wild-type and mutant-LNA probes that hybridize at the mutation site. The fourplex real-time RT-PCR assay using LNA-based TaqMan probes exploits the 5’-3’ nuclease activity of Taq DNA polymerase, allowing direct detection of each of the four fluorogenic qPCR products by the uncoupling of a reporter dye from its quencher dye during qPCR. Furthermore, LNA modification of TaqMan probes increases their base pairing stability and creates a highly favorable context of duplex formation between them and their target sequences under more stringent conditions (Braasch and Corey 2001; McTigue et al. 2004; Petersen and Wengel 2003; Ugozzoli et al. 2004). Therefore, these properties allow LNA-based probes to discriminate between distinct viral genomic sequences that differ by a single nucleotide at a precise hot spot mutational site. This was performed, in three different places on the genome of SARS-CoV-2 at the same time into one reaction. In this way, it is possible to detect a range of representative genotypes that could reveal the presence of different variants of SARS-CoV-2 geographically. This method for differential diagnosis of SARS-CoV-2 virus isolates could be of help for effective surveillance. Furthermore, this multitarget rRT-PCR assay enables rapid follow-up of the most clinically relevant variants across the population in real-time and effectively guide the selection of viral strains that disserve further analysis by their whole-genome sequencing using next-generation sequencing methods.

## Data Availability

All data are included in the present manuscript. Oligonucleotides, gBlocks and plasmids used for this study are available if requested. The authors state that all results obtained for confirming the conclusions presented in the article are represented fully within the article.

## Acknowledgments

We are grateful to Dr. Gilles Dupuis for critical reading of the manuscript and for his valuable comments. We thank Dr. Hugues Charest for his communication with respect to the N LSPQ primer-probe set and its utilization in rRT-PCR assays. This study was supported by the Ministère de l’Économie et de l’Innovation (MÉI) and, in part, by the Natural Sciences and Engineering Research Council of Canada (NSERC, grant #ALLRP 554545-20) to S.Labbé.

## Author contributions

M.D. conceptualized, performed and analyzed rRT-PCR data. P.T. conceptualized the design of primers and TaqMan-LNA probes as well as carrying numerous bioinformatic analyses. S. Lévesque conceptualized and performed BioFire FilmArray assays. A.B. acquired and analyzed the data. A.C, L.V., and P.M. conceptualized the experimental work and analyzed data. S. Labbé conceptualized the experimental work and wrote the manuscript. The authors reviewed the results and approved the final version of the manuscript.

## Footnotes

^1^ Abbreviations used are: B2M, β_2_-microglobulin; C_t_, threshold cycle; cDNA, complementary DNA; LNA, locked nuclei acid; PCR, polymerase chain reaction; NATRVP, NATtrol Respiratory verification panel; RT, reverse transcription; rRT-PCR, real-time quantitative RT-PCR; SNP, single nucleotide polymorphism.

## Supplemental figure legend

**Fig. S1.**
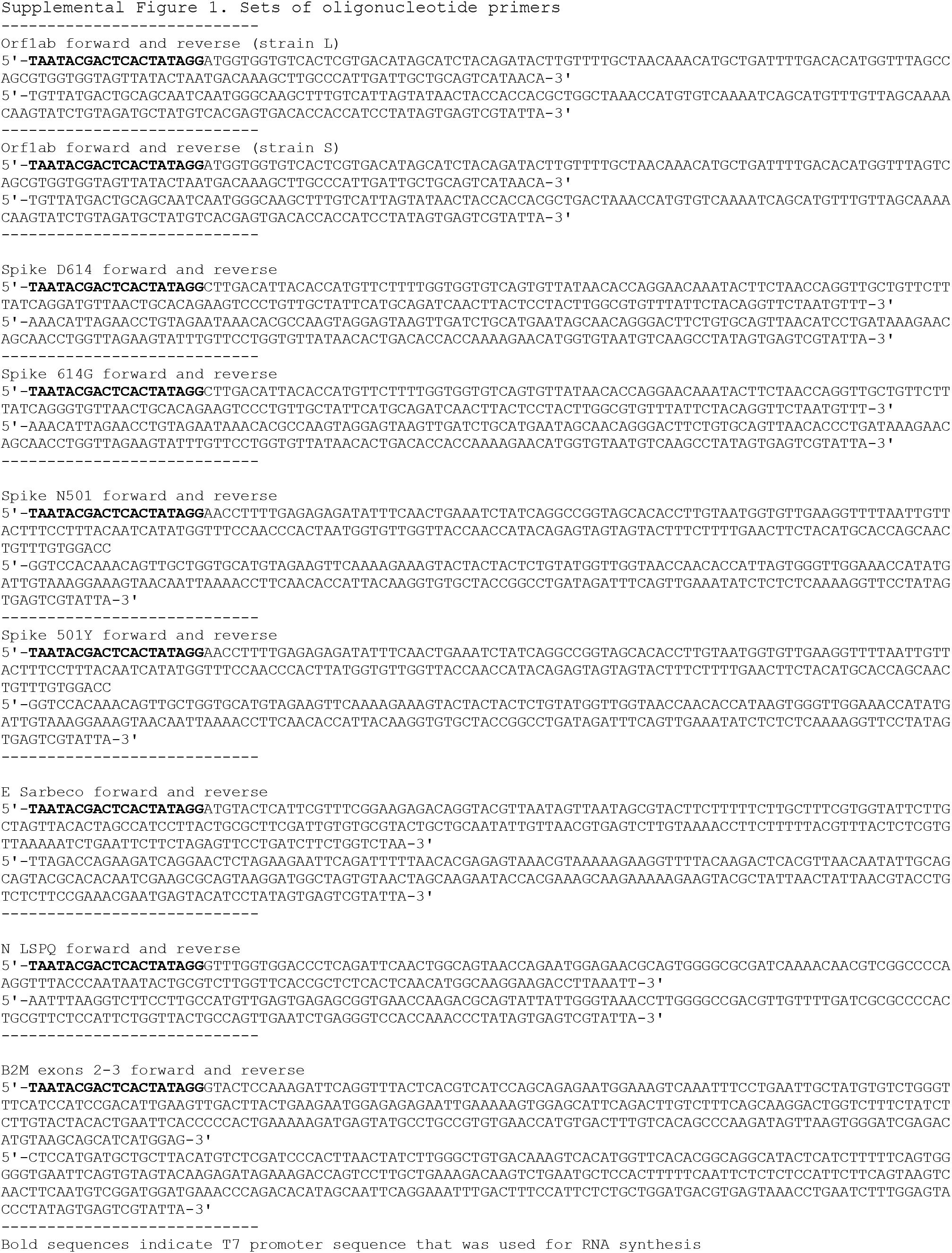
Sequences of the synthetic oligomers used to create the indicated gBlocks. Each targeted SARS-CoV-2 region from the Orf1ab. Spike. E. and N genes (according to the Wuhan-Hu-1 genome reference sequence and mutant derivatives) were designed accompanied with a T7 promoter sequence (bold) at their 5’ends.

